# Face mask use and associated factors among students in rural Eastern Uganda amidst the COVID-19 pandemic

**DOI:** 10.1101/2021.06.27.21259131

**Authors:** Denis Mwesige, Aisha Nalugya, Douglas Bulafu, Arnold Tigaiza, Bridget Tamale Nagawa, Emmanuel Balinda, Abel Wilson Walekhwa

## Abstract

**Background:** The Corona Virus Disease of 2019 (COVID-19) has gravely affected several aspects of national and global society, including education. Given the risk it poses, the Government of Uganda (GOU) adopted and recommended face mask use as one of the preventive measures to limit its transmission in communities. However, there is limited data on the levels of face mask usage and associated factors among students in schools in Uganda. This study aimed at assessing the face mask usage and associated factors among students in schools in rural Eastern Uganda amidst the COVID-19 pandemic.

**Methods:** A cross sectional quantitative descriptive study was conducted among 423 students in schools in rural Eastern Uganda. Multi-stage sampling method was employed in the selection of study participants. The data was collected by trained data collectors using structured questionnaires pre-installed on ODK enabled smart phones. The data entered was cleaned using Excel 2016 and exported to Stata14.0 statistical software (Statacorp, College station, Texas, USA) for analysis. Bivariate and multivariable logistic regression analyses were employed using 95% CI (confidence interval). Variables with *p-*value < 0.20 and those with literature backup evidence were included in the multivariable model. Variables with *p-*value < 0.05 were considered to be statistically significant. This study revealed that less than three quarters (62.3%) wore face masks correctly.

**Results:** Almost all, 98.9% of the participants mentioned that they wore face masks due to fear of missing classes and 49.0% disagreed that they were vulnerable to COVID-19. Students in boarding schools (AOR = 1.61, 95%CI: 1.05-2.47), those who believed that they were vulnerable to COVID-19 (AOR = 1.70, 95%CI: 1.11-2.10), and those who disagreed that masks are uncomfortable (AOR = 1.62, 95%CI: 1.06-2.46) were more likely to wear facemasks correctly.

**Conclusion:** This study revealed that more than a third of the students did not wear face masks correctly. Correct wearing of face masks was associated with being in a boarding school, belief that they were susceptible to COVID-19, and disagreeing that masks were uncomfortable. This therefore highlights the need for sensitization programmes in academic institutions in order to improve students’ perceptions toward COVID-19 and face masks, and consequently increase correct face mask usage in schools.

## Background

Globally, Severe Acute Respiratory Syndrome coronavirus 2 (SARS-CoV-2), the causative agent of COVID-19 has spread to several countries and territories, tremendously impacting health, education, trade, transport, and economies (McKibbin and Fernando, 2020). As of 22nd June 2021, approximately 178,202,610 confirmed cases and 3,865,738 COVID-19 related deaths had been reported globally (WHO, 2021). Situation analyses by Uganda’s ministry of health (MOH) indicated a surge in the number of cases, with 72,679 confirmed cases and 680 deaths recorded as of 22^nd^ June (MOH, 2021). The COVID-19 pandemic has affected all spheres of life with a huge effect realized on educational institutions from pre-primary to tertiary levels (Adeshakin et al., 2020, El-Sadr and Justman, 2020). A total of 948 cases had been reported in 43 schools in 22 districts in Uganda as of 6^th^ June 2021 (MOH, 2021). However, this is an underestimate of the actual number of cases in schools due to underreporting.

The first case of COVID-19 in Uganda was recorded on 21^st^ March 2020. In an attempt to limit its spread, the Government of Uganda instituted several measures, including a nationwide lockdown, closure of border points of entry, mandatory quarantine(Mboowa et al., 2020), and closure of all education institutions to protect the 15 million learners at the different levels (MOH, 2020). In addition, measures such as hand hygiene, social distancing and use of face masks were adopted and recommended (MOH, 2020, Mboowa et al., 2020). On 20^th^ September 2020, the president of Uganda announced the reopening of education institutions for candidate classes and final year students at higher institutions of learning. Subsequently, all classes reopened in a phased manner from March to mid-June 2021 (MOES, 2020). The Ministry of Education and Sports (MOES), together with the MOH and other stakeholders issued guidelines and standard operating procedures (SOPs) for creation of a safe environment for learners and staff amidst the COVID-19 pandemic (MOES, 2020). Despite the existence of these guidelines, a considerable number of infection clusters have been reported in schools in Uganda (MOH, 2021).

The World Health Organization (WHO) currently recommends medical-grade mask for people over 60 years when they are out and cannot socially distance, while the general public is advised to wear a non-medical mask (three-layer fabric mask) (WHO, 2020). Besides, appropriate use, storage and cleaning or disposal of masks is essential to ensure that they are as effective as possible and to avoid an increased transmission risk (WHO, 2020, Mboowa et al., 2021). In Uganda, the Government supported expansion of local manufacturing of masks with a plan to distribute at least one free mask to all individuals aged 6 years and older (Nannyonga et al., 2020). However, the usage of face masks among several communities in Uganda, including schools is still unknown.

Existing studies around the world have focused on face mask use among university students (Barrios et al., 2021, Duong et al., 2021), with little attention given to primary and secondary students. Yet, schools have become sources of infection clusters in many countries, including Uganda. Therefore, this study aimed at assessing use of face masks and associated factors among primary and secondary students in rural Eastern Uganda in order to inform interventions targeted at improving face mask use, and consequently reduce the risk of transmission of COVID-19 in schools.

## Methods

### Study design and setting

A cross sectional study utilizing quantitative data collection methods was conducted during the period of March-April 2021 after partial re-opening of schools in Uganda. This study was conducted in Tirinyi Town Council found in Kibuku District Eastern Uganda. Kibuku District has fourteen sub counties and five town councils (urban areas) of Kibuku Town council (TC), Bulangira TC, Tirinyi TC, Kasasira TC, and Kadama TC. Kibuku district is boarded by Pallisa District in the North, Budaka in the East, Butaleja district in the South and Namutumba District in the West. Kibuku district has a total of 402 villages. Tirinyi town council has both government and private schools, primary and secondary schools, but no tertiary institution.

### Sample size and sampling procedure

The sample size was estimated using the Kish Leslie formula for cross-sectional studies (Kish, 1965). A prevalence (p) of facemask use in Uganda was taken to be 50% owing to the fact that there were limited studies conducted on facemask use. A 95% level of confidence, an error rate (d) of 0.05 and a Z score of 1.96 corresponding to the two 95% confidence interval (CI) and a 10% non-response were used in the calculation. This yielded a sample size of 423.

Multistage sampling technique was used. A list of all the functional schools in the town council was obtained from the office of District Education Officer. We then used random sampling to select ten schools from the list using the Ms Excel randomizer. Private and public schools were included in this study to ensure inclusion of students from all socio-economic strata. The study recruited students aged 10 years and above, in primary and secondary schools. Therefore, sampling frames of students aged 10 years and above, were obtained from each selected school. A random sample of students was then selected using Ms Excel randomizer.

### Data collection technique and tools

Interviews with students were conducted using a structured questionnaire in order to assess face mask use and associated factors. Data were also collected through observing the practices of facemask use in the schools. The questionnaire was pre-installed on the open data kit (ODK) on the smart phones of the research assistants.

### Study variables and measurement

The dependent variable for our study was “correct use of face masks”. The dependent variable was categorized as Yes/No. A respondent was considered to be correctly using a face mask if they were wearing it over the nose and mouth, all the way to the chin. The independent variables included socio-demographics such as age of student, sex, school type (boarding vs. day) and class level. Other independent variables included, attitude and knowledge variables.

### Data management and analysis

Data were collected and entered using the Open data kit (ODK), and synchronized onto the server daily. Mobile data collection allowed for real-time data capture and entry and minimized errors throughout the data management process. Data entry screens were designed with skips and restrictions to ensure quality and completeness. Only the study team had access to the data. Data were then exported for analysis in Stata 14.0 statistical software (Statacorp, College station, Texas, USA). Initially, simpler regression models consisting of the outcome and one predictor at a time were run to produce unadjusted odds ratios. Variables with p values less than 0.2 in the bivariable models and those with literature backup evidence were included in the multivariable model. A p-value ≤ 0.05 was considered statistically significant.

### Quality control and assurance

Research assistants with vast experience in research were recruited in this study. Prior to data collection, we trained research assistants for two days on the data collection tool, ethics and study protocol. The study tools were pretested at Kirika Sub-county, one of the sub counties in Kibuku district in order to ensure familiarity of the research assistants with the tool and also identify any errors. During data collection, research assistants were supervised to ensure that they followed the study protocol when conducting observations and interviewing respondents. All investigators and research assistants undertook an online course on good clinical practice for a better understanding of human’s research.

## Results

### Socio-demographic characteristics of the students in rural eastern Uganda

Nearly three quarters, 73.8% (312/423) of the participants were 10-15 years old, 55.3% (234/423) were male and more than a third, 37.2% (157/423) were Anglican. More than half, 64.8% (274/423) of the participants were in day schools and 51.5% (218/423) were in P.5-P7 classes (Table 1).

**Table 1:** Socio-demographic characteristics of the students in rural eastern Uganda

| Variable | Frequency (n=423) | Percentage (%) |
| --- | --- | --- |
| **Age** |  |  |
| 10-15 years | 312 | 73.8 |
| 16-20 years | 106 | 25.1 |
| 21 and above | 5 | 1.2 |
| **Sex** |  |  |
| Male | 234 | 55.3 |
| Female | 189 | 44.7 |
| **School type** |  |  |
| Boarding | 149 | 35.2 |
| Day | 274 | 64.8 |
| **Class level** |  |  |
| P.4 and below | 59 | 14.0 |
| P.5 - P.7 | 218 | 51.5 |
| S.1 - S.4 | 135 | 31.9 |
| S.5 - S.6 | 11 | 2.6 |
| **Religion** |  |  |
| Anglican | 157 | 37.2 |
| Catholic | 92 | 21.8 |
| Moslem | 86 | 20.3 |
| Pentecostal | 71 | 16.8 |
| Seventh Day Adventist | 12 | 2.8 |
| Others | 4 | 1.0 |

### Face-mask use among primary and secondary school students in rural eastern Uganda

Majority, 89.6% (379/423) of the participants owned a face mask. Of these, 90.0% (341/379) had cotton cloth masks given to them by the government of Uganda (Table 2). However, more than a third, 37.7% (143/379) did not wear the face masks correctly at the time of the interview. Almost all, 98.9% (375/379) of the study participants only wore face masks due to fear of missing classes, while 1.1% (4/379) wore them due to fear of COVID-19. (Table 2)

**Table 2:**
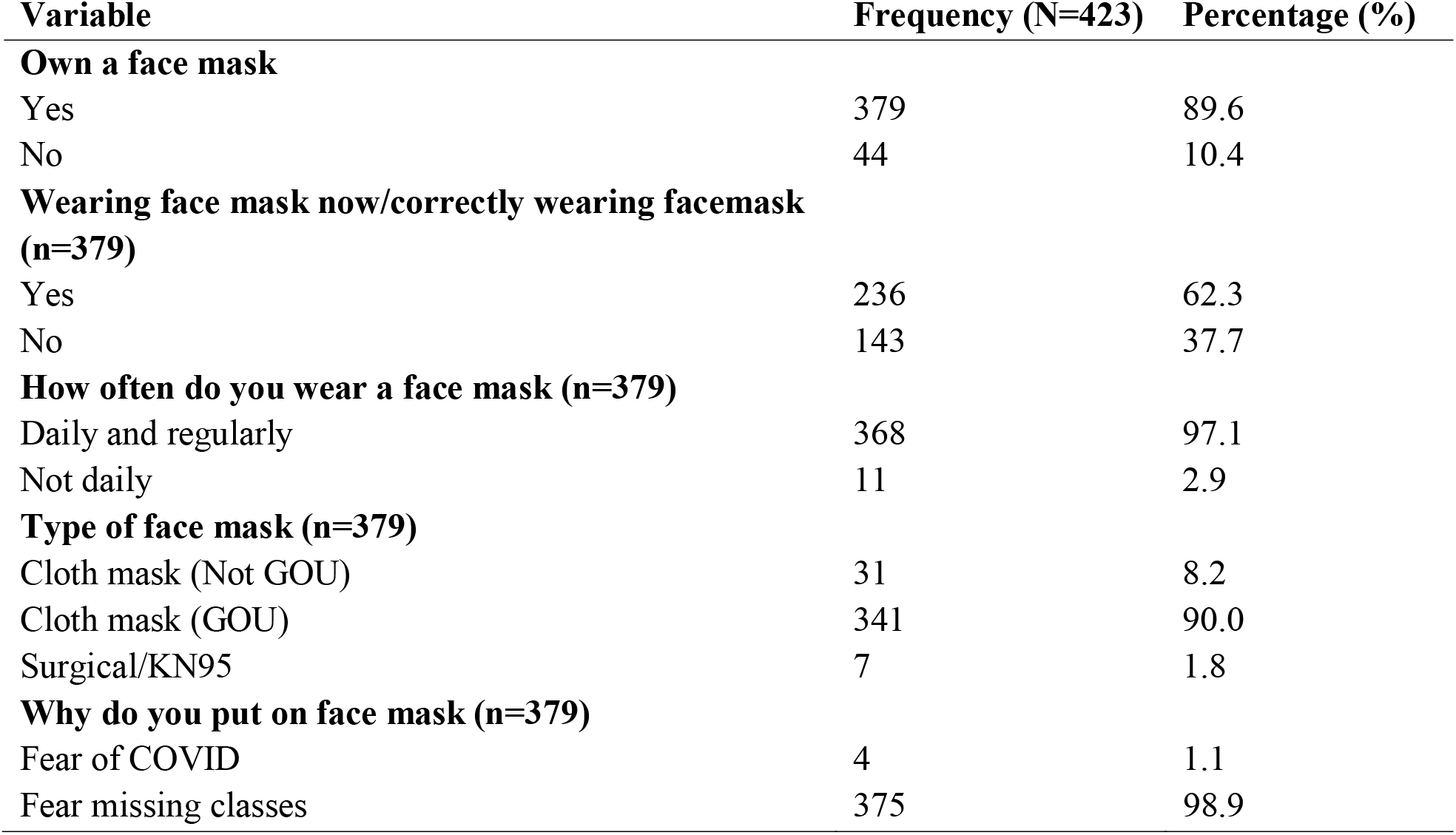
Face mask use among students in rural Eastern Uganda

### Perceptions towards Face-mask use and COVID-19 among primary and secondary school students in rural Eastern Uganda

Nearly half, 49.0% (205/423) of the participants disagreed that were vulnerable to COVID-19 disease. Majority, 91.2% (386/423) of the participants agreed that appropriate size mask is important, 94.3% agreed that a mask should cover the nose, mouth and chin, and 96.0% believed that it’s important to wash and iron a face mask. More than half, 58.2% (246/423) agreed that face masks are uncomfortable. (Table 3)

**Table 3:** Students’ perceptions toward COVID-19 and face masks in rural Eastern Uganda

| **Variable** | **Frequency (N=423)** | **Percentage (%)** |
| --- | --- | --- |
| **Think that they are vulnerable to COVID 19** |  |  |
| Yes | 195 | 46.7 |
| No | 205 | 49.0 |
| Maybe | 18 | 4.3 |
| **Appropriate size of mask is important** |  |  |
| Agree | 386 | 91.2 |
| Disagree | 37 | 8.8 |
| **Mask should cover the nose, mouth and chin** |  |  |
| Agree | 399 | 94.3 |
| Disagree | 24 | 5.7 |
| **It's important to wash and iron a facemask** |  |  |
| Agree | 406 | 96.0 |
| Disagree | 17 | 4.0 |
| **Face masks are uncomfortable** |  |  |
| Agree | 246 | 58.2 |
| Disagree | 177 | 41.8 |
| **Wear mask because teacher wears mask** |  |  |
| Agree | 162 | 38.3 |
| Disagree | 261 | 61.7 |

### Factors associated with Face-mask use among primary and secondary school students in Uganda

At bivariate analysis, being male (AOR=1.61, 95%CI=1.06-2.44), P<0.05), being in a boarding school (AOR=1.61, 95%CI=1.05-2.47), p<0.05), belief of being vulnerable to COVID-19 (AOR=1.70, 95%CI=1.11-2.10, p<0.05), and disagreeing that masks are uncomfortable (AOR=1.62, 1.06-2.46), P<0.05) was positively associated with proper wearing of face-masks.

After adjusting for confounders, students in boarding sections/schools (AOR=1.61, 95%CI=1.05-2.47), p<0.05), were 1.61 times more likely to wear masks properly compared to those in day schools/sections. Those who agreed that they were vulnerable to COVID-19 (AOR=1.70, 95%CI=1.11-2.10), p<0.05), were 1.70 times more likely to wear masks properly compared to those who thought they were not vulnerable. Students that disagreed that masks are uncomfortable (AOR=1.62, 95%CI=1.06-2.46), P<0.05), were 1.62 times more likely to wear masks properly compared to those who agreed that masks were uncomfortable (Table 4)

**Table 4:** Factors associated with face mask use among primary and secondary school students in rural Eastern Uganda

| Variables | Correctly wearing facemasks (%) |  | COR(95% CI) | AOR(95% CI) |
| --- | --- | --- | --- | --- |
|  | No (N=236) | Yes (N=143) |  |  |
| **Age** |  |  |  |  |
| 10-15 years | 76.3 | 73.4 | 1 |  |
| 16-20 years | 22.5 | 25.9 | 1.20 (0.74-1.94) |  |
| 21 and above | 1.2 | 0.7 | 0.57 (0.06-5.56) |  |
| **Sex** |  |  |  |  |
| Male | 59.3 | 47.6 | **1.61(1.06-2.44)\*** | 1.43(0.92-2.23) |
| Female | 40.7 | 52.4 | 1 | 1 |
| **School type** |  |  |  |  |
| Boarding | 32.2 | 43.4 | **1.61(1.05-2.47)\*** | **1.59 (1.03-2.44)\*** |
| Day | 67.8 | 56.6 | 1 | 1 |
| **Class level** |  |  |  |  |
| P.4 and below | 13.6 | 16.8 | 1 |  |
| P.5 - P.7 | 52.1 | 49.7 | 0.77 (0.420-1.409) |  |
| S.1 - S.4 | 32.2 | 29.4 | 0.74 (0.385-1.411) |  |
| S.5 - S.6 | 2.1 | 4.2 | 1.60 (0.436-5.868) |  |
| **Religion** |  |  |  |  |
| Anglican | 36.0 | 38.5 | 1 |  |
| Catholics | 21.6 | 19.6 | 0.85 (0.48-1.50) |  |
| Moslems | 21.2 | 20.3 | 0.89 (0.51-1.58) |  |
| Pentecostals | 16.5 | 18.2 | 1.03 (0.57-1.88) |  |
| SDA | 4.2 | 1.4 | 0.31 (0.65-1.46) |  |
| Others | 0.4 | 2.1 | 4.64 (0.47-45.71) |  |
| **Type of face mask (n=379)** |  |  |  |  |
| Cloth mask (Not GOU) | 8.5 | 7.7 | 1 |  |
| Cloth mask (GOU) | 89.4 | 90.9 | 1.12(0.52-2.41) |  |
| Surgical/KN95 | 2.1 | 1.4 | 0.73(0.12-4.39) |  |
| **Why do you put on face mask (n=379)** |  |  |  |  |
| Fear of COVID | 98.7 | 99.3 | 1.83(0.19-17.75) |  |
| Fear missing classes | 1.27 | 0.7 | 1 |  |
| **Think that they are vulnerable to COVID 19** |  |  |  |  |
| Yes | 41.8 | 55.9 | **1.70 (1.11-2.10)\*** | **1.68 (1.08-2.61)\*** |
| No | 53.5 | 42.0 | 1 | 1 |
| Maybe | 4.7 | 2.1 | 0.56 (0.15-2.10) | 0.60 (0.16-2.31) |
| **Appropriate size of mask is important** |  |  |  |  |
| Agree | 91.5 | 93.7 | 1 |  |
| Disagree | 8.5 | 6.3 | 0.73 (0.32-1.64) |  |
| **Mask should cover the nose, mouth and chin** |  |  |  |  |
| Agree | 95.3 | 94.4 | 1 |  |
| Disagree | 4.7 | 5.6 | 1.21 (0.48-3.09) |  |
| **It's important to wash and iron a facemask** |  |  |  |  |
| Agree | 91.5 | 93.7 | 1 |  |
| Disagree | 8.5 | 6.3 | 0.40 (0.11-1.44) |  |
| **Face masks are uncomfortable** |  |  |  |  |
| Agree | 61.4 | 49.6 | 1 | 1 |
| Disagree | 38.6 | 50.4 | **1.62 (1.06-2.46)\*** | **1.60 (1.04-2.47)\*** |
| **Wear mask because teacher wears mask** |  |  |  |  |
| Agree | 39.4 | 38.5 | 1 |  |
| Disagree | 60.6 | 61.5 | 1.04(0.68-1.59) |  |

## Discussion

Despite the reported increase in the clusters of infections in Uganda’s education institutions which later led to closures, there is limited literature on facemask use amongst students. This study aimed at assessing facemask use and associated factors among students in schools in rural Eastern Uganda during the COVID-19 pandemic. The study showed that more than a third of the respondents were not correctly wearing facemasks at the time of interview. Nearly half, of the respondents thought that they were not vulnerable to COVID-19 disease. Furthermore, students in boarding sections/schools, those who perceived being vulnerable to COVID-19 and those who disagreed with masks being uncomfortable were more likely to wear them correctly compared to their counterparts.

This study revealed that less than three quarters (62.3%) of the respondents wore facemasks correctly at the time of interview. These findings don’t concur with those in a cross-sectional study conducted by Barrios and colleagues which revealed that 89.7% of the interviewed students were wearing facemasks correctly (Barrios et al., 2021). The difference in study findings could be attributed to the level of education of the students and the difference in study settings. Barrios and colleagues conducted their study among university students in the United States who undoubtedly have better access to information and are therefore more knowledgeable about the risk of COVID-19 infection and the importance of wearing facemasks unlike those in rural eastern Uganda (Barrios et al., 2021). Furthermore, our study findings could be explained by the low COVID-19 risk perception reported in this study. Close to half (49%) of the students in this study believed that they were not vulnerable to COVID-19. These findings imply that there’s need for strict enforcement of face mask usage by school administrations, so as to change students’ behavior. Furthermore, continuous sensitizations on correct face mask use should be done.

The study also revealed that majority of the masks being worn by students were reusable cloth masks. Our findings are in agreement with those by Barrios et al. (2021) in USA which found that 92.2% of the students wore reusable cloth masks. The findings however differ from those in a study conducted by Duong et al. (2021) in Vietnam which revealed that only 23.1% were using reusable cloth masks. Reusable cloth masks have been recommended for use among the general public, including in schools by the WHO and Uganda’s ministry of health (WHO, 2020), which explains the usage levels among students. In addition, the government of Uganda distributed cloth masks to its population as part of the preventive interventions to limit the spread of COVID-19 (Nannyonga et al., 2020). Besides, cloth masks are re-useable, durable and hence cheaper in the long run for students in rural Eastern Uganda who might not be able to afford a new mask every other day. However, the discrepancy with results reported by Duong could be due to the perceived high risk for COVID-19 infection since they were experiencing continuous domestic outbreaks in Vietnam at the time (Vietnam, 2020) therefore the choice of surgical masks (Duong et al., 2021). Given the significant number of students using reusable cloth masks in this study, it’s important to emphasize proper storage and cleanliness so as to ensure that the masks are as effective as possible in reducing transmission of COVID-19.

It’s worth noting that nearly half of the respondents thought they were not vulnerable to COVID-19. This is quite unfortunate but true and could be attributed to the poor access to information especially in rural areas. Students could also have felt that they were not vulnerable to COVID-19 since the rural areas where they reside have generally been perceived as safe zones and urban areas as high risk areas. This perception could impact correct face mask usage and uptake of other SOPs. However, our findings don’t corroborate those in studies conducted in south Western Ethiopia (53.4%), Ghana (68.3%) and Sierra Leone (75%) (Asefa et al., 2020, Serwaa et al., 2020, Tadese et al., 2021) which indicated higher risk perceptions. Our findings show the need for more efforts toward sensitization of students in rural areas on their risk and susceptibility to COVID-19.

At multivariable analysis, students who perceived that they were vulnerable to COVID-19 were more likely to wear facemasks correctly. This is not surprising because a high risk perception increases the need to take up prevention measures in an attempt to preserve one’s health. These findings corroborate those by Sim and colleagues who reported that individuals are more likely to wear facemasks due to the perceived susceptibility and perceived severity of being afflicted with life-threatening diseases (Sim et al., 2014). Risk perceptions and health behavior have also been associated in several other studies (Ferrer and Klein, 2015, Brewer, 2007). Respondents who disagreed with the statement that masks were uncomfortable were also more likely to wear them correctly. Facemasks are one of the recommended preventive measures in controlling the transmission of COVID-19 and comfort of a facemask is one the factors that determine its appropriate use. Sim and colleagues reported in their literature review that personal discomfort was one of the perceived barriers to proper facemask use which resonates with our finding. In light of this, it is important to promote facemask use among other standard operating procedures in order to control the spread of COVID-19. Students in boarding sections/schools were statistically more likely to wear masks correctly compared with those in day sections/schools. This can be attributed to the fact that teachers and staff in boarding schools could be strictly enforcing COVID-19 measures compared to day schools, since students are left under complete care of the school administration. As a result, the school administration might feel more obligated to keep students safe.

## Strengths and limitations

This is one of the few studies that has established the prevalence of face mask usage among students in Uganda amidst the COVID-19 pandemic. The study used a relatively large sample size, making the results more reliable. The study employed observational methods to assess correctness of facemask wearing. Therefore, this could have brought about observer bias. In case the students were aware that they were being observed, it could have caused them to act differently from how they normally would. This was limited by ensuring that research assistants were well trained. We also did not collect data on knowledge which could probably have informed COVID-19 risk perceptions of the students. We recommend more studies on; Adherence to facemask use among students, teachers and lecturers in education institutions including qualitative studies to understand the salient issues that could explain the adherence levels.

## Conclusion

Our study revealed that more than a third did not correctly use face masks. Nearly half of the students did not believe that they were at risk for COVID-19 and this is quite scary as it could influence adherence to facemask use as well as other SOPs. Being in a boarding sections/schools, perception of vulnerability to COVID-19 and finding masks comfortable were associated with a higher likelihood of wearing facemasks correctly. Interventions aimed at increasing knowledge on risk factors for COVID-19, changing perceptions and promoting adherence to SOPs in schools are therefore warranted to ensure safety of students in such populous environments.

## Data Availability

The data that support the findings of this study are available from the corresponding author upon request.

## Authors’ contributions

All authors reviewed and approved the final manuscript. DM and AWW conceptualized the study, drafted the study protocol, participated in data collection and analysis and drafted the manuscript. AN, DB, AT, BNT and EB participated in the analysis and drafting of the manuscript.

## Ethical Considerations

Ethical approval was obtained from the Cavendish University Uganda Research and ethics committee. Permission was also obtained from Kibuku district local government leadership prior to the data collection. Administrative clearance was also sought from all schools from which participants were selected. In addition, informed consent was obtained from all research participants. Consent from a parent/ guardian was sought for participants below 18 years of age. Participants aged 10-17 years were asked to provide their own assent in addition to the parental consent form. Unique identifiers were used to protect the identity of the participants involved in the study. Access to the collected data was restricted to the study team.

